# Impact of intensive care-related factors on outcome in stroke patients, results from the population-based Brest Stroke Registry: an observational study

**DOI:** 10.1101/2023.05.17.23290128

**Authors:** Philippe Ariès, Pierre Bailly, Thibaut Baudic, Fanny Le Garrec, Maëlys Consigny, Erwan L’Her, Serge Timsit, Olivier Huet the Brest Stroke Registry collaborators

## Abstract

**Background:** Little is known on the burden of ICU care for stroke patients. The aim of this study was to provide a description of management strategies, resource use, complications and their association with prognosis of stroke patients admitted to ICU.

**Methods:** Using a population-based stroke registry, we analyzed consecutive stroke patients admitted to 3 ICU with at least one organ failure between 2008 and 2017. The study period was divided into two periods corresponding to the arrival of mechanical reperfusion technique.

Predictors of ICU mortality were separately assessed in two multivariable logistic regression models, a “clinical model” and an “intervention model”. The same analysis was performed for predictors of functional status at hospital discharge.

**Results:** 215 patients were included. Stroke etiology was ischemia in 109 patients (50.7%) and hemorrhage in 106 patients (49.3%). Median NIHSS score was 20.0 (9.0; 40.0). The most common reason for ICU admission was coma (41.2%) followed by acute circulatory failure (41%) and respiratory failure (27.4%). 112 patients (52%) died in the ICU and 20 patients (11.2%) had a good functional outcome (mRS≤3) at hospital discharge.

In the “clinical model,” factors independently associated with ICU mortality were: age (OR = 1.03 [95%CI, 1.0 to 1.06]; *p=0.04*) and intracranial hypertension (OR = 6.89 [95%CI, 3.55 to 13.38]; *p<0.0001*). In the “intervention model,” the need for invasive mechanical ventilation (OR = 7.39 [95%CI, 1.93 to 28.23]; *p=0.004*), the need for vasopressor therapy (OR = 3.36 [95%CI, 1.5 to 7.53]; *p=0.003*) and decision of withholding life support treatments (OR = 19.24 [95%CI, 7.6 to 48.65]; *p<0.0001*) were associated with bad outcome.

**Conclusion:** Our study showed the very poor prognosis of acute stroke patients admitted to ICU. These results also suggest that the clinical evolution of these patients during ICU hospitalization may provide important information for prognostication.

## Background

Prognosis of stroke patients admitted into the intensive care unit (ICU) remains very poor despite stroke unit’s care and recent advances in acute reperfusion treatment for ischemic strokes (1). The reported short-term mortality rate for stroke patients requiring ICU management ranged from 40 to 70% (2–6) and the likelihood of survival without disabilities was very low in a recent study, since only a fifth of patients had a good neurological outcome at 6 months after stroke (2).

For these reasons, a time-limited-trial (TLT) of intensive care treatment-i.e., agreement between physicians and family to use certain medical therapies over a defined period (7)-may be proposed (8). Yet, despite TLT, decision of invasive therapy and treatment of complications might be considered as futile by the care-team and could lead to a self-fulfilling prophecy in patients who could have, if treated aggressively, a reasonable outcome (9). Therefore, ICU management plays a key role in the prognosis of critically ill stroke patients but little is known about how intensive care-related factors may impact prognosis contrary to pre-hospital anamnesis (8,10–12).

The need for mechanical ventilation is associated with poor outcomes but data concerning other life-supporting interventions such as the need of vasopressor support or renal replacement therapy are scarce (5). Moreover, the impact of stroke-related complications such as intracranial hypertension, status epilepticus and neurosurgery on ICU stroke patients’ outcomes remains poorly investigated.

In addition, the negative effect of systemic secondary brain insults (SBIs): hypoxaemia, hypercapnia, hypotension, fever, anemia, hyperglycemia and abnormal blood sodium levels, on cerebral blood flow, as well as oxygen and glucose supply have been less well characterized in critically ill stroke patients contrary to other acute brain injuries such as traumatic brain injury (13,14).

Thus, the primary objective of this study was to determine critical care-related factors associated with ICU mortality in patients enrolled in the French Brest Stroke Registry (BSR), a stroke population-based registry set up in 2008 in western France, an area that includes about 336 000 individuals (15–17). We also aimed to determine factors associated with functional outcome at hospital discharge.

## Methods

### Study population

Our study concerned all strokes from the BSR which occurred between January 2008 and December 2017. All cases of stroke in patients aged above 15 years occurring in a defined area known as the “Pays de Brest”, in Brittany, western France, were recorded in the BSR. The total population (above 15 years) of this area was 365 564 at the 2009 census. Data collection was performed prospectively and retrospectively (“hot and cold pursuit”). Multiple information sources were used: emergency wards, brain imaging records, neurologists and general practitioners, death certificates and hospital database. The World Health Organization (WHO) criteria (18) were used to make the clinical diagnosis of stroke, confirmed by brain CT or MRI. Stroke was defined as a new focal neurological deficit with symptoms and signs persisting for more than 24 hours (patients who died within the first 24 hours were also included), or a focal neurological deficit lasting at least one hour, or resolving in less than one hour but presenting with brain imaging (CT or MRI) suggestive of stroke.

### Criteria of inclusion

All patients admitted in the ICU with a diagnosis of ischemic or hemorrhagic stroke and registered in the BSR with at least one organ failure among circulatory failure, respiratory failure and neurological failure were included.

Acute circulatory failure was defined as a persistent hypotension (mean arterial pressure (MAP) below 65 mmHg) requiring vasopressors or inotropes associated or not with hyperlactatemia (> 2mmol/l), respiratory failure as the need for high flow nasal cannula oxygen therapy (HFNC) and/or non-invasive ventilation (NIV) and/or IMV for respiratory reasons, and neurological failure as coma i.e., a Glasgow Coma Scale (GCS) of 7 or less.

Patients could be hospitalized in one of the three ICUs (one neurosurgical, two general ICUs) of the corresponding geographic area of the BSR. We excluded patients without hospitalization reports or in case of patient’s refusal to participate.

### Ethics & specific authorizations

BSR is accredited by the French National Agency of Health surveillance (Santé Publique France) and complies with the French regulation on patient’s consent, ethics, and data confidentiality. Specific authorizations were obtained from the national “Comité consultatif sur le traitement de l’information en matière de recherche” under the reference CCTIRS MG/CP°07.693 and from the “Commission Nationale Informatique et Liberté” (CNIL) under the agreement N° 908085. The local ethic committee also approved the registry.

The study protocol was submitted to the Institutional Review Board of the Brest University Hospital (B2020CE.17) and was registered on ClinicalTrials.gov public website (NCT04434287). Patients or relatives were individually informed by postal mail and had the opportunity to decline participation, but no written informed consent was required, in line with the directives of the Ethics Committee.

### Data collection

Data prospectively collected in the BSR included risk factors of stroke and the initial clinical examination (modified Rankin score (mRS) before stroke, National Institutes of Health Stroke Scale (NIHSS) and GCS at hospital admission). Specific variables according to the type of stroke were also collected, i.e., for ischemic strokes: the trial of ORG 10172 in acute stroke treatment (TOAST) classification, the Oxfordshire Community Stroke Project (OSCP) classification, radiological findings (stroke localization) and management specificities (thrombolysis, thrombectomy for e.g.); for hemorrhagic strokes: specific etiologies and radiological findings (stroke localization, imaging progression) (19–21).

We also retrospectively collected specific data related to the ICU hospitalization:

- Data from admission: the Simplified Acute Physiology Score II (SAPS II), the type of organ failure and the presence of aspiration pneumonia was also collected;
- Systemic secondary brain insults (SBIs) during the first 24 hours: hypoxemia (oxygen saturation (SatO2) <92% or partial arterial pressure of oxygen (PaO2) <8 kPa without oxygen-therapy or mechanical ventilation with FIO2 > 30% for respiratory reason), hypercapnia (partial pressure of carbon dioxide (PaCO2) > 6 kPa), hypotension (systolic blood pressure (SBP) below 90 mmHg and/or use of vasopressors and/or inotropes to maintain SBP above 90 mmHg), hyperglycemia (glucose ≥ 8 mmol/l without insulin therapy), hyperthermia (temperature ≥ 38° Celsius), anemia (serum hemoglobin < 10g/dl before transfusion), hyponatremia (natremia <135 mmol/l), hypernatremia (natremia >155 mmol/l). The physiologic values were measured every 4 hours in the ICU. Blood samples were usually performed every morning. One deviation during the 24 hours was sufficient to be recorded;
- Stroke-related complications during the ICU stay: intracranial hypertension (ICH) defined either by intracranial pressure (ICP) above 20 mmHg during more than 30 minutes in case of ICP monitoring, or by an abnormal trans-cranial doppler (TCD) measures i.e., pulsatility index above 1.4 or diastolic flow velocity under 20 cm.s-1 without hypocapnia, or clinically by unilateral or bilateral mydriasis requiring medical or surgical intervention (such as sedation, osmotherapy or neurosurgical intervention), status epilepticus and neurosurgery (i.e., craniectomy, hematoma evacuation, external ventricular drain);
- Organ support during the ICU stay: need for IMV, and/or for NIV and/or for HFNC oxygen; the use of vasopressor support (i.e., noradrenalin, adrenalin or dobutamine) or for antihypertensive therapy and/or for renal replacement therapy (RRT). For ventilated patients, the indication for IMV was also assessed;
- Other complications during the ICU stay: Ventilator Acquired Pneumonia (VAP) were defined as a pneumonia occurring >48 hours after endotracheal intubation with clinical and radiological signs associated with bacterial documentation. Early-onset and late-onset VAP were respectively defined as VAP occurring between day 2 and 6 and after day 7 of ICU admission (22). Radiological proven deep venous thrombosis or pulmonary embolism, duration of invasive ventilation, ICU length of stay, withholding and withdrawal of life-sustaining treatments (LST) were also collected;
- Specific monitoring: invasive intracranial pressure (ICP) monitoring, TCD, and cardiac output (CO) monitoring (invasive or not monitoring).

### Outcomes

We assessed the ICU mortality, the primary outcome, and the functional status (mRS) at hospital discharge. According to previous studies, we defined a good hospital discharge functional outcome as a mRS score of 0 to 3 (0, no disability; 1, no significant disability; 2, slight disability; and 3,moderate disability) (4,12) and bad outcome (mRS >3).

In order to analyze temporal trends in these outcomes, we divided the study period into two periods: 2008-2013 and 2014-2017 corresponding to the arrival of endovascular mechanical reperfusion technique in Brest University Hospital, which may have an impact on outcomes.

### Statistical Analysis

Characteristics of patients were described as frequencies and percentages for categorical variables, as means and standard deviations (SD) or medians and interquartile ranges (IQR) for continuous variables. Categorical variables were analyzed by chi-square test or Fisher’s exact test and continuous variables were compared by Student’s t test or by Wilcoxon rank test according to their distribution.

We built two different models for each outcome, a “clinical model” with demographic, clinical examination, biological and radiological variables and a “intervention model” including organ supports and withholding or withdrawal of LST.

Considering the rule that predictive logistic models should be used with a minimum of 10 outcome events per predictor variable, we estimated that a number of 200 included patients would be sufficient to propose logistic regression models with 5 variables as mortality for ICU stroke patients was estimated to be 40% (11). The initial neurologic gravity (NIHSS and GCS score) and decision of withholding LST were considered as important factors and were forced respectively in the clinical and the intervention model to be tested.

The set of variables associated with ICU survival and good functional prognosis in univariate analysis (p<0.05) were included in a multivariate regression models and backward stepwise selection was applied. The variance inflation factor (VIF) was applied to detect the presence of collinearity between independent variables of multivariable models. No variable included in models had missing data greater than 10% therefore we did not perform imputation and all available data have been analyzed.

Tests were two-sided and values of *p* less than 0.05 were considered statistically significant. Statistical analysis was performed with SAS 9.4 (SAS Institute Inc.,Cary, NC, USA).

## Results

### Patients enrolled

Out of 8910 patients included in the Brest Stroke Registry during the study period, 338 patients were hospitalized in ICU and 215 were finally enrolled **(Figure 1)**.

**Fig.1.**
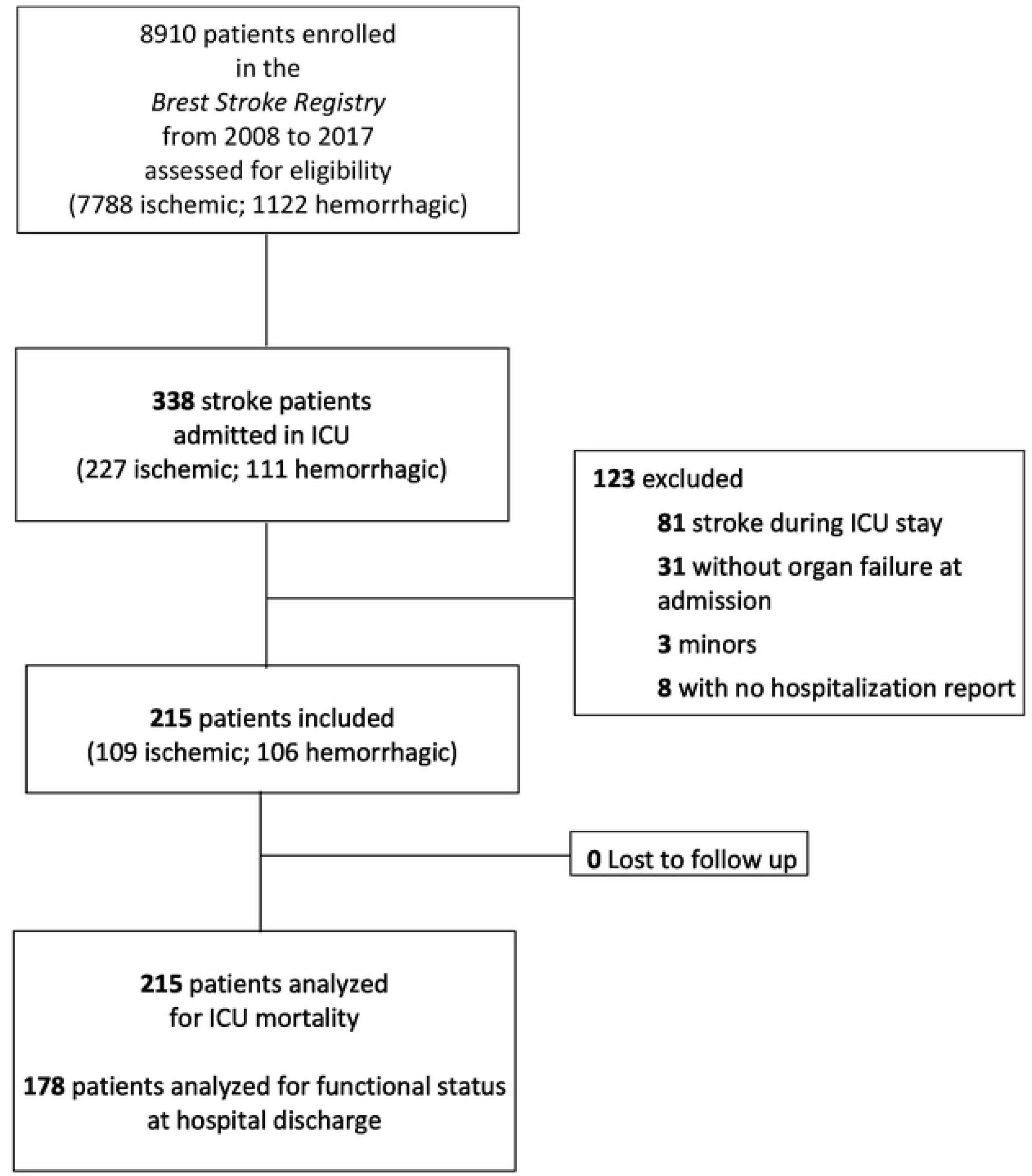
Flowchart. *ICU: Intensive Care Unit*

One hundred and twenty-eight patients were enrolled from 2008 to 2013 and 87 from 2014 to 2017. Ischemic stroke was diagnosed in 109/215 (50.7 %) patients and ICH occurred in 106/215 (49.3 %) patients. Patients were predominantly males (61.4%), median age was 66.0 (57.0-76.0) years, median SAPS II score was 55.0 (43.0-67.0) and median NIHSS was 20.0 (9.0- 40.0). The most frequent organ failure at admission was coma (41.2%), followed by acute circulatory failure (41%) and respiratory failure (27.4%). One hundred and twelve (52%) patients died in ICU and median mRS at hospital discharge was 4.0 (3.0-6.0).

Only 20 patients (11.2%) had a good functional outcome (mRS≤3) **(Figure 2)**.

**Fig.2.**
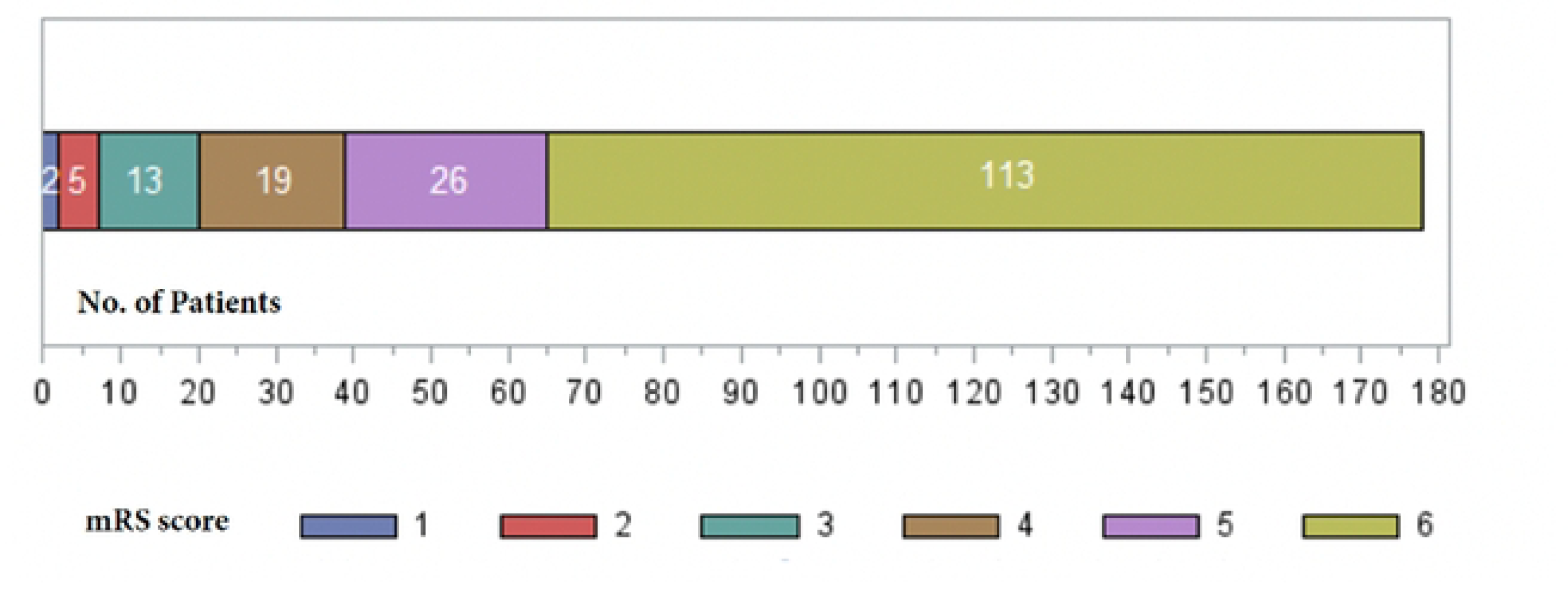
Modified Rankin Scale at Hospital discharge. *mRS: modified Rankin Scale*

### Univariate and multivariable analysis for ICU mortality

In univariate analysis, admission factors associated with ICU-mortality were age, SAPS II, NIHSS, GCS score, TOAST and OCSP classifications **(Table 1)**.

**Table 1.**
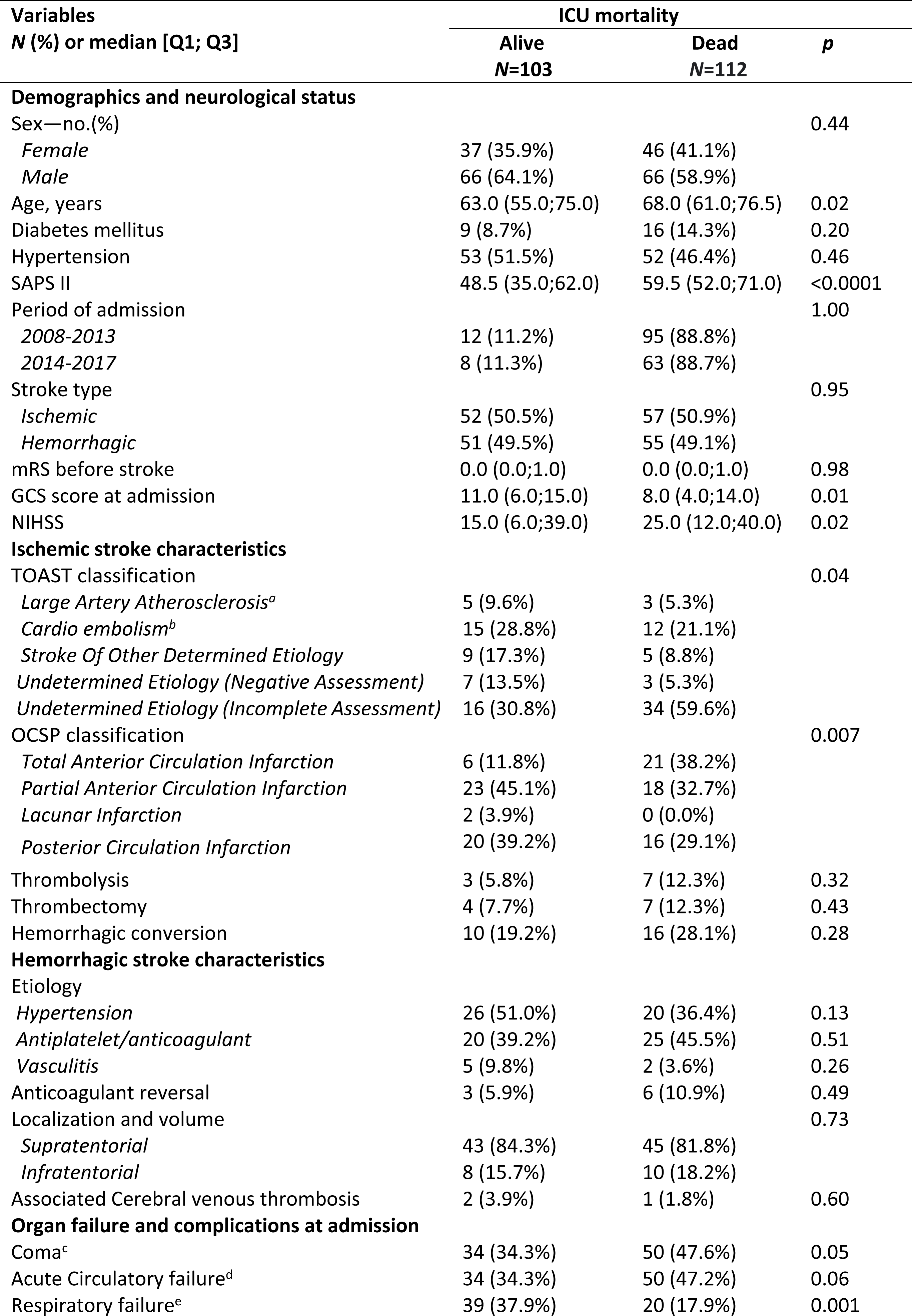
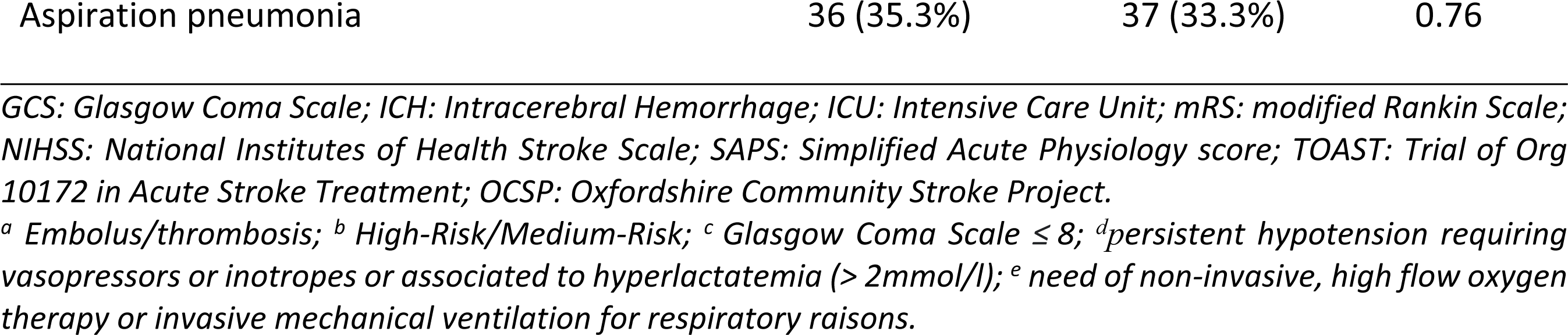
Admission characteristics and their univariate association with ICU mortality.

Among critical care-related factors, respiratory failure, invasive mechanical ventilation, vasopressor use and the occurrence of intracranial hypertension were associated with ICU mortality. During the ICU stay, hyperglycemia and hypernatremia were the only systemic SBIs associated with mortality. ICU length of stay was significantly longer in the survivor group compared to those who died in ICU (8 days (4.0;20.0) *vs* 3 days (2.0;6.0), p<0.001) **(Table 2)**.

**Table 2.** Critical care-related factors and their univariate association with ICU mortality.

| Variables<br>N (%) or median [Q1; Q3] | ICU mortality |  | p |
| --- | --- | --- | --- |
|  | Alive<br>N=103 | Dead<br>N=112 |  |
| **Systemic secondary brain insults during the first 24 hours** |  |  |  |
| Hypoxemia<sup>a</sup> | 33 (34.4%) | 30 (28.8%) | 0.40 |
| Hypercapnia<sup>b</sup> | 29 (30.9%) | 40 (40.4%) | 0.17 |
| Hypotension<sup>c</sup> | 34 (34.3%) | 50 (47.2%) | 0.06 |
| Hyperglycemia<sup>d</sup> | 19 (20.4%) | 35 (35.0%) | 0.02 |
| Hyperthermia<sup>e</sup> | 47 (46.1%) | 36 (34.3%) | 0.08 |
| Anemia<sup>f</sup> | 26 (27.1%) | 18 (16.7%) | 0.07 |
| Hyponatremia<sup>g</sup> | 34 (34.3%) | 23 (21.7%) | 0.04 |
| Hypernatremia<sup>h</sup> | 2 (2.0%) | 15 (14.2%) | 0.0016 |
| **Complications during ICU stay** |  |  |  |
| Intracranial hypertension | 21 (20.4%) | 73 (65.8%) | <0.0001 |
| Status epilepticus | 23 (22.3%) | 7 (6.4%) | 0.0008 |
| Neurosurgery<sup>i</sup> | 32 (31.1%) | 21 (18.9%) | 0.04 |
| Imaging progression<sup>j</sup> | 10 (20.8%) | 14 (53.8%) | 0.004 |
| VAP | 16 (15.7%) | 11 (9.9%) | 0.21 |
| Timing of VAP |  |  | 0.82 |
| *Early onset pneumonia* | 8 (50.0%) | 6 (54.5%) |  |
| *Late onset pneumonia* | 8 (50.0%) | 5 (45.5%) |  |
| Thrombosis<sup>k</sup> | 8 (7.8%) | 5 (4.5%) | 0.32 |
| **Organ support during ICU stay** |  |  |  |
| Invasive mechanical ventilation | 69 (67.0%) | 107 (95.5%) | <0.0001 |
| Reasons for mechanical ventilation |  |  | 0.91 |
| *Coma* | 54 (79.4%) | 90 (84.1%) |  |
| *Respiratory failure* | 12 (17.6%) | 14 (13%) |  |
| *Elective procedure* | 2 (2.9%) | 3 (2.8%) |  |
| Oxygen trough NIV or High flow nasal cannula | 31 (30.4%) | 8 (7.2%) | <0.0001 |
| Use of vasopressors | 43 (42.6%) | 78 (70.9%) | <0.0001 |
| Use of antihypertensive treatments | 54 (52.9%) | 27 (25.2%) | <0.0001 |
| Use of renal replacement therapy | 5 (4.9%) | 8 (7.3%) | 0.46 |
| **Specific monitoring** |  |  |  |
| Cardiac monitoring | 23 (22.3%) | 30 (26.8%) | 0.45 |
| TCD monitoring<sup>l</sup> | 32 (31%) | 25 (22%) | 0.32 |
| Invasive ICP monitoring | 6 (5%) | 5 (4%) | 0.51 |
| **Outcomes** |  |  |  |
| Duration of invasive ventilation, days | 4.0 (0.0;11.0) | 3.0 (2.0;6.0) | 0.99 |
| GCS at discharge | 15.0 (13.0;15.0) | 3.0 (3.0;3.0) | <0.0001 |
| mRS at discharge | 4.0 (3.0;6.0) | 6.0 (6.0;6.0) | <0.0001 |
| ICU length of stay, days | 8.0 (4.0;20.0) | 3.0 (2.0;6.0) | <0.0001 |
| Withholding LST | 9 (8.7%) | 70 (63.1%) | <0.0001 |
| Withdrawal LST | 3 (2.9%) | 56 (50.5%) | <0.0001 |

GCS: Glasgow Coma Scale; ICP: Intracranial Pressure; ICU: Intensive Care Unit; LST: Life- Sustaining Treatment; mRS: modified Rankin Scale; NIV: Non-Invasive Ventilation; TCD: Transcranial doppler; VAP: Ventilation Acquired Pneumonia. <sup>a</sup> SatO<sub>2</sub> <92% or partial arterial pressure of oxygen (PaO<sub>2</sub>) <8 or FIO<sub>2</sub> > 30% kPa; <sup>b</sup> PaCO<sub>2</sub> > 6 kPa; <sup>c</sup> vasopressors or inotropes to maintain MAP>60mmHg; <sup>d</sup> glucose≥8 mmol/l; <sup>e</sup> temperature ≥ 38° Celsius; <sup>f</sup> serum haemoglobin<10g/dl; <sup>g</sup> Na+<135 mmol/l; <sup>h</sup> Na+>155 mmol/l, <sup>i</sup> craniectomy, hematoma evacuation, external ventricular drain; <sup>j</sup> increase in hemorrhage volume from baseline CT scan; <sup>k</sup> proven venous thrombosis or pulmonary embolism; <sup>l</sup> At least one TCD measure during the ICU stay

The multivariate analysis for ICU mortality is presented in **Table 3**. The clinical model showed that age and intracranial hypertension were independent clinical risk factors associated with ICU mortality. The intervention model showed that IMV, use of vasopressors and withholding of LST were independently associated with ICU mortality. In contrast, hyponatremia, status epilepticus and oxygen trough NIV or HFNC were the only variables associated with survival.

**Table 3.** Multivariable Logistic Regression of clinical factors interventions associated with ICU mortality.

| Variable | Full model |  | Final model |  |
| --- | --- | --- | --- | --- |
|  | OR [95%CI] | *p* | OR [95%CI] | *p* |
| Age | 1.02 [0.99 ; 1.06] | 0.22 | 1.03 [1.00 ; 1.06] | 0.04 |
| NIHSS score at admission | 1.02 [0.98 ; 1.07] | 0.34 |  |  |
| GCS score at admission | 1.04 [0.89 ; 1.20] | 0.65 |  |  |
| Hyperglycemia<sup>a</sup> | 2.08 [0.78 ; 5.55] | 0.14 |  |  |
| Hyponatremia<sup>b</sup> | 0.45 [0.17 ; 1.18] | 0.10 | 0.48 [0.23 ; 0.98] | 0.04 |
| Intracranial hypertension | 5.94 [2.48 ; 14.21] | <0.0001 | 6.89 [3.55 ; 13.38] | <0.0001 |
| Status epilepticus | 0.22 [0.04 ; 1.28] | 0.09 | 0.34 [0.13 ; 0.92] | 0.03 |
| Respiratory failure<sup>c</sup> | 0.25 [0.08 ; 0.81] | 0.02 |  |  |

**(B) Intervention model**
| Variable | Full model |  | Final model |  |
| --- | --- | --- | --- | --- |
|  | OR [95%CI] | *p* | OR [95%CI] | *p* |
| Invasive mechanical ventilation | 6.68 [1.79 ; 24.96] | 0.005 | 7.39 [1.93 ; 28.23] | 0.004 |
| Use of vasopressors | 3.49 [1.52 ; 8.01] | 0.003 | 3.36 [1.50 ; 7.53] | 0.003 |
| Use of antihypertensive treatments | 0.53 [0.23 ; 1.24] | 0.16 |  |  |
| Oxygen trough NIV or High flow nasal cannula | 0.25 [0.08 ; 0.82] | 0.02 | 0.20 [0.06 ; 0.60] | 0.004 |
| Neurosurgery<sup>d</sup> | 0.52 [0.20 ; 1.33] | 0.17 |  |  |
| Withholding LST | 20.01 [7.70 ; 52.03] | <0.0001 | 19.24 [7.60 ; 48.65] | <0.0001 |

*CI: Confidence Interval; LST: Life- Sustaining Treatment; NIV: Non-Invasive Ventilation; OR: Odds Ratio.*
<sup>a</sup>glucose ≥ 8 mmol/l; <sup>b</sup> Na+ < 135 mmol/l; <sup>c</sup> SatO2 < 92% or partial arterial pressure of oxygen (PaO2) < 8 or FIO2 >
30% kPa; , <sup>d</sup> craniectomy, hematoma evacuation, external ventricular drain

Variance Inflation Factors were small (below 1.5) suggesting very low correlation between regression variables **(Table 4)**.

**Table 4.** Multicollinearity analysis using Variance Inflation Factor (VIF)

| Variables | VIF |
| --- | --- |
| **ICU mortality** |  |
| ***Clinical model*** |  |
| Age | 1.04 |
| Hyponatremia | 1.01 |
| Intracranial hypertension | 1.03 |
| Status epilepticus | 1.07 |
| ***Intervention model*** |  |
| Invasive mechanical ventilation | 1.19 |
| Use of vasopressors | 1.14 |
| Oxygen trough NIV or High flow nasal cannula | 1.05 |
| Withholding LST | 1.08 |

*ICU: Intensive Care Unit; LST: Life- Sustaining Treatment; NIV: Non-Invasive Ventilation, VIF: Variance Inflation Factor.*

### Univariate and multivariable analysis for functional outcome at hospital discharge

In univariate analysis, variables associated with a poor functional outcome at hospital discharge (mRS ≥ 4) were age, SAPS II score, ischemic stroke, NIHSS score, coma at admission, the need of IMV and of vasopressors. Occurrence of hyperglycemia and intracranial hypertension were also associated with a poor functional outcome **(Table 5).**

**Table 5.**
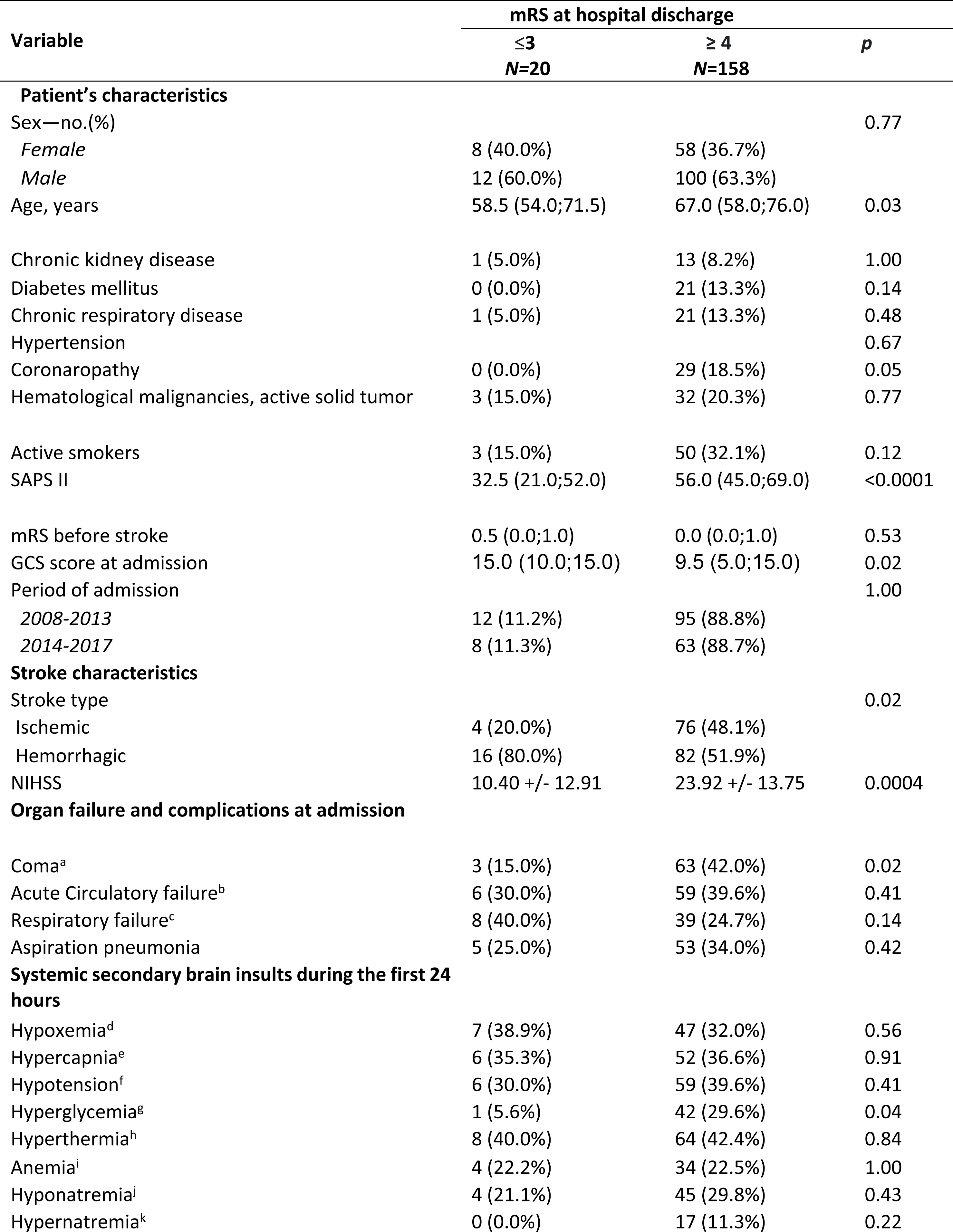
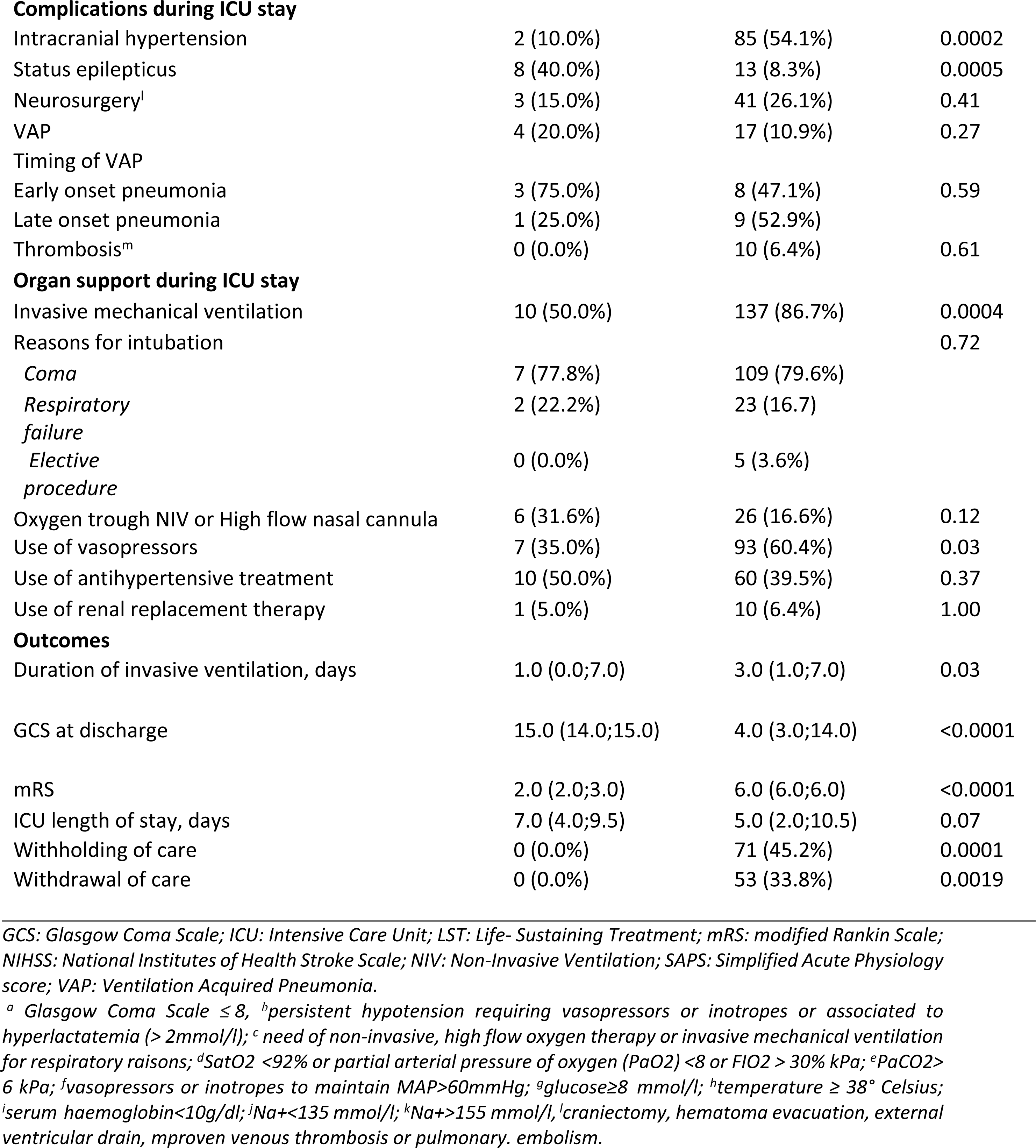
Baseline characteristics, ICU management, outcomes and their univariate association with functional prognosis at hospital discharge.

| Variable | mRS at hospital discharge |  | *p* |
| --- | --- | --- | --- |
|  | ≤3<br>*N*=20 | ≥ 4<br>*N*=158 |  |
| **Patient's characteristics** |  |  |  |
| Sex—no.(%) |  |  | 0.77 |
| *Female* | 8 (40.0%) | 58 (36.7%) |  |
| *Male* | 12 (60.0%) | 100 (63.3%) |  |
| Age, years | 58.5 (54.0;71.5) | 67.0 (58.0;76.0) | 0.03 |
| Chronic kidney disease | 1 (5.0%) | 13 (8.2%) | 1.00 |
| Diabetes mellitus | 0 (0.0%) | 21 (13.3%) | 0.14 |
| Chronic respiratory disease | 1 (5.0%) | 21 (13.3%) | 0.48 |
| Hypertension |  |  | 0.67 |
| Coronaropathy | 0 (0.0%) | 29 (18.5%) | 0.05 |
| Hematological malignancies, active solid tumor | 3 (15.0%) | 32 (20.3%) | 0.77 |
| Active smokers | 3 (15.0%) | 50 (32.1%) | 0.12 |
| SAPS II | 32.5 (21.0;52.0) | 56.0 (45.0;69.0) | <0.0001 |
| mRS before stroke | 0.5 (0.0;1.0) | 0.0 (0.0;1.0) | 0.53 |
| GCS score at admission | 15.0 (10.0;15.0) | 9.5 (5.0;15.0) | 0.02 |
| Period of admission |  |  | 1.00 |
| 2008-2013 | 12 (11.2%) | 95 (88.8%) |  |
| 2014-2017 | 8 (11.3%) | 63 (88.7%) |  |
| **Stroke characteristics** |  |  |  |
| Stroke type |  |  | 0.02 |
| Ischemic | 4 (20.0%) | 76 (48.1%) |  |
| Hemorrhagic | 16 (80.0%) | 82 (51.9%) |  |
| NIHSS | 10.40 +/- 12.91 | 23.92 +/- 13.75 | 0.0004 |
| **Organ failure and complications at admission** |  |  |  |
| Coma<sup>a</sup> | 3 (15.0%) | 63 (42.0%) | 0.02 |
| Acute Circulatory failure<sup>b</sup> | 6 (30.0%) | 59 (39.6%) | 0.41 |
| Respiratory failure<sup>c</sup> | 8 (40.0%) | 39 (24.7%) | 0.14 |
| Aspiration pneumonia | 5 (25.0%) | 53 (34.0%) | 0.42 |
| **Systemic secondary brain insults during the first 24 hours** |  |  |  |
| Hypoxemia<sup>d</sup> | 7 (38.9%) | 47 (32.0%) | 0.56 |
| Hypercapnia<sup>e</sup> | 6 (35.3%) | 52 (36.6%) | 0.91 |
| Hypotension<sup>f</sup> | 6 (30.0%) | 59 (39.6%) | 0.41 |
| Hyperglycemia<sup>g</sup> | 1 (5.6%) | 42 (29.6%) | 0.04 |
| Hyperthermia<sup>h</sup> | 8 (40.0%) | 64 (42.4%) | 0.84 |
| Anemia<sup>i</sup> | 4 (22.2%) | 34 (22.5%) | 1.00 |
| Hyponatremia<sup>j</sup> | 4 (21.1%) | 45 (29.8%) | 0.43 |
| Hypernatremia<sup>k</sup> | 0 (0.0%) | 17 (11.3%) | 0.22 |
| **Complications during ICU stay** |  |  |  |
| Intracranial hypertension | 2 (10.0%) | 85 (54.1%) | 0.0002 |
| Status epilepticus | 8 (40.0%) | 13 (8.3%) | 0.0005 |
| Neurosurgery<sup>l</sup> | 3 (15.0%) | 41 (26.1%) | 0.41 |
| VAP | 4 (20.0%) | 17 (10.9%) | 0.27 |
| Timing of VAP |  |  |  |
| Early onset pneumonia | 3 (75.0%) | 8 (47.1%) | 0.59 |
| Late onset pneumonia | 1 (25.0%) | 9 (52.9%) |  |
| Thrombosis<sup>m</sup> | 0 (0.0%) | 10 (6.4%) | 0.61 |
| **Organ support during ICU stay** |  |  |  |
| Invasive mechanical ventilation | 10 (50.0%) | 137 (86.7%) | 0.0004 |
| Reasons for intubation |  |  | 0.72 |
| Coma | 7 (77.8%) | 109 (79.6%) |  |
| Respiratory failure | 2 (22.2%) | 23 (16.7%) |  |
| Elective procedure | 0 (0.0%) | 5 (3.6%) |  |
| Oxygen trough NIV or High flow nasal cannula | 6 (31.6%) | 26 (16.6%) | 0.12 |
| Use of vasopressors | 7 (35.0%) | 93 (60.4%) | 0.03 |
| Use of antihypertensive treatment | 10 (50.0%) | 60 (39.5%) | 0.37 |
| Use of renal replacement therapy | 1 (5.0%) | 10 (6.4%) | 1.00 |
| **Outcomes** |  |  |  |
| Duration of invasive ventilation, days | 1.0 (0.0;7.0) | 3.0 (1.0;7.0) | 0.03 |
| GCS at discharge | 15.0 (14.0;15.0) | 4.0 (3.0;14.0) | <0.0001 |
| mRS | 2.0 (2.0;3.0) | 6.0 (6.0;6.0) | <0.0001 |
| ICU length of stay, days | 7.0 (4.0;9.5) | 5.0 (2.0;10.5) | 0.07 |
| Withholding of care | 0 (0.0%) | 71 (45.2%) | 0.0001 |
| Withdrawal of care | 0 (0.0%) | 53 (33.8%) | 0.0019 |

GCS: Glasgow Coma Scale; ICU: Intensive Care Unit; LST: Life-Sustaining Treatment; mRS: modified Rankin Scale; NIHSS: National Institutes of Health Stroke Scale; NIV: Non-Invasive Ventilation; SAPS: Simplified Acute Physiology score; VAP: Ventilation Acquired Pneumonia.
<sup>a</sup> Glasgow Coma Scale $\leq 8$ , <sup>b</sup> persistent hypotension requiring vasopressors or inotropes or associated to hyperlactatemia ( $> 2\text{ mmol/l}$ ); <sup>c</sup> need of non-invasive, high flow oxygen therapy or invasive mechanical ventilation for respiratory reasons; <sup>d</sup> SatO<sub>2</sub> $< 92\%$ or partial arterial pressure of oxygen (PaO<sub>2</sub>) $< 8$ or FIO<sub>2</sub> $> 30\%$ kPa; <sup>e</sup> PaCO<sub>2</sub> $> 6$ kPa; <sup>f</sup> vasopressors or inotropes to maintain MAP $> 60\text{ mmHg}$ ; <sup>g</sup> glucose $\geq 8$ mmol/l; <sup>h</sup> temperature $\geq 38^\circ$ Celsius; <sup>i</sup> serum haemoglobin $< 10\text{ g/dl}$ ; <sup>j</sup> Na<sup>+</sup> $< 135$ mmol/l; <sup>k</sup> Na<sup>+</sup> $> 155$ mmol/l, <sup>l</sup> craniectomy, hematoma evacuation, external ventricular drain, <sup>m</sup> proven venous thrombosis or pulmonary embolism.

Multivariate analysis of functional outcome at hospital discharge is presented in **Table 6**. The clinical model showed that high NIHSS at admission was associated with poor outcomes contrary to high GCS score and status epilepticus that were associated with a good functional outcome. Invasive mechanical ventilation was the only variable of the intervention model associated with poor functional outcome at hospital discharge.

**Table 6.** Multivariate Logistic Regression of clinical factors and intervention associated with functional prognosis at hospital discharge.

| Variables | OR | 95%CI | *p* |
| --- | --- | --- | --- |
| **Clinical model** |  |  |  |
| NIHSS at admission (per point) | 1.07 | [1.00 ; 1.14] | 0.05 |
| GCS at discharge (per point) | 0.53 | [0.29 ; 0.96] | 0.04 |
| Status epilepticus | 0.10 | [0.01 ; 0.83] | 0.03 |
| **Intervention model** |  |  |  |
| Invasive mechanical ventilation | 6.52 | [2.43 ; 17.55] | 0.0002 |

*CI: Confidence Interval; GCS: Glasgow Coma Scale; NIHSS: National Institutes of Health Stroke Scale; OR: Odds Ratio.*

## Discussion

Our study showed that the prognosis of the 215 stroke patients admitted to ICU with organ failures was poor with an ICU mortality of 52% and only 11% of the patients surviving had a good functional outcome at hospital discharge. We identified strong associations between critical-care-related factors and outcomes. Indeed, the occurrence of intracranial hypertension and the need for organ support therapy (IMV and vasopressor therapy) were independent predictors of ICU mortality. Among these parameters, only the need for IMV was independently associated with a poor functional outcome at hospital discharge.

The strongest independent clinical predictor for ICU death was the occurrence of ICH during the ICU stay. Using a standardized definition, we observed that this complication was frequent as it occurred in 43.4% of patients. This finding is in line with previous reports and suggests that critically ill stroke patients died primarily from stroke-related complications occurring during the ICU stay (2,11,23). However, the use of invasive ICP monitoring was very low as only 5% of patients were monitored. Compared to TBI patients, the monitoring of ICP in stroke patients remains poorly investigated and is not routinely recommended in patients with coma after hemorrhagic or ischemic stroke (24,25). There is evidence that ICP can be elevated even following decompressive hemicraniectomy or after hematoma removing (26,27).

In contrast, status epilepticus during the ICU stay, another stroke-related complication, was associated with a better outcome. It has been proposed that this protective effect was related to the reversibility of the initial coma when status epilepticus was the etiology of the neurologic failure (23).

The strong association between some of the organ support therapies and outcome found in our study provides new insights into the comprehension of the impact of acute phase therapies on prognosis. Our intervention model identified that both IMV and vasopressor support were independent predictors of ICU death contrary to NIV or HFNC oxygen therapy. As the majority of patients required intubation because of coma for airway management (82.3%) and vasopressor therapy is usually used for patients who developed intracranial hypertension in order to maintain the cerebral blood perfusion, we hypothesized that IMV and vasopressors mainly reflected the severity of the stroke. Another main hypothesis to explain this association relies on the burden of care in ICU. Indeed, life-supporting therapies as IMV or vasopressor support are associated with numerous adverse events and iatrogenic consequences that, in turn, may worsen outcomes.(28)

An interesting finding is the absence of impact of systemic SBIs on outcome. The secondary brain damages refer to delayed functional or structural damages observed after various types of acute cerebral injuries. A large part of these damages is due to systemic SBIs such as hypotension, hypoxemia or fever that increase the neurological damage by preventing the regulation of cerebral blood flow (13,29). As systemic SBIs are physiologic abnormalities which can be easily treated, it represents the cornerstone for severe TBI management (29). Moreover, a large part of these pathomechanisms is supposed to be common between the different acute cerebral injuries (30).

For non-ICU stroke patients, arterial hypotension is rare, but SBP under 100 mmHg or diastolic blood pressure under 70 mmHg has been associated with poor neurological outcomes (31). However, it remains unclear whether systemic SBIs impact outcomes for ICU stroke patients. Our results suggest that contrary to TBI or SHA patients (29,32), systemic SBI may not be deleterious. Recently, Fontaine *et al.* found similar results for critically ill patients with convulsive status epilepticus (CSE) (33). The ongoing multicenter study SPICE that aims to identify the effect of major systemic complications will help to precise the impact of SBIs on outcome (12).

In contrast, the association between hyponatremia and ICU survival could be explain by the higher probability to develop hyponatremia during the ICU stay for survivors compared to patients who died. Indeed, the ICU length of stay was significantly longer in the survivor group compared to those who died in ICU.

Except for age, all independent predictors of the ICU mortality for critically ill stroke patients were related to the ICU stay (i.e., life support therapies and complications), contrary to the neurologic severity scores at admission as NIHSS score or Glasgow coma scale.

These results suggest that ICU management strategy (i.e.: resources used) could impact on survival. Despite a global poor prognosis, some patients may benefit of an ICU hospitalization and have a good functional recovery. Therefore, it has been proposed to consider long enough TLT for these patients (34) as a too short TLT may expose to inappropriate limitation of organ supports and to a self-fulfilling prophecy. In contrast, a too long TLT may expose to a potential loss of compassion and decreased quality of care (7). Our study did not evaluate the good duration of TLT, but we consider that it should be long enough to evaluate specific indicators, as it has been proposed for elderly patients admitted to ICU (35). For ICU stroke patients these indicators could be the need or not of IMV and vasopressor support and the occurrence or not of intracranial hypertension.

Our study has some limitations. First, the BSR registry has not been designed for ICU studies and contrary to stroke specific data, all ICU data have been retrospectively recorded.

Second, we used the “ten events per variable (EPV) rule” to determine the number of subjects required although current evidence supporting for binary logistic regression is weak (36). Third, although the BSR could provide long-term mortality, we couldn’t study long-term functional status. Fourth, the observational design of the study did not allow us to explore causality. Finally, due to the high rate of withholding LST, we could not avoid the bias of self- fulfilling prophecy.

## Conclusion

Despite a poor global prognosis, a part of stroke patients with organ failures may benefit from ICU admission and life-sustaining interventions. Therefore, prognostication is key but still remains challenging. We found that occurrence of intracranial hypertension and the need of IMV or vasopressor support were strong predictors of ICU mortality. Our study suggests that critical care-related factors could provide valuable information for prognostication in addition to the initial neurological severity. Our findings warrant further studies to evaluate the impact of ICU care (notably SBI prevention and treatment) on patient’s prognostication.

## Data Availability

All relevant data are within the manuscript and its Supporting Information files.

## Acknowledgements

We would like to thank the Brest Stroke Registry collaborators:

Philippe Goas*, Irina Viakhireva-Dovganyuk*, François Mathias Merrien*, Aurore Jourdain*, François Rouhart*, Amélie Leblanc*, Marie Bruguet*, Denis Marechal*, Jordan Coris* *CHRU Brest, Department of Neurology & Stroke Unit, CHRU de Brest, Université de Bretagne Occidentale, Brest, 29200, France.

